# Lower perceived social support is significantly associated with elevated levels of psychological distress in racially and ethnically diverse close family members of cardiac arrest survivors

**DOI:** 10.1101/2024.02.25.24303342

**Authors:** Mina Yuan, Isabella M Tincher, Bhanvi Sachdeva, Sabine Abukhadra, Danielle A Rojas, Christine DeForge, Sachin Agarwal

## Abstract

**Background:** Poor perceived social support has been associated with worse psychological distress in close family members after their loved one’s hospitalization with prolonged mechanical ventilation, but never been tested after cardiac arrest.

**Methods:** Close family members of consecutive cardiac arrest patients hospitalized at an academic tertiary care center were recruited before hospital discharge, and perceived social support was assessed using the Multidimensional Scale of Perceived Social Support (MSPSS). Indicators of psychological distress were administered via telephone at 1 month after cardiac arrest. Multivariate linear regressions were used to estimate the associations between MSPSS total score and total Patient Health Questionnaire-8 (PHQ-8) score (primary outcome) and total PTSD (PCL-5) and generalized anxiety (GAD-2) scores, after adjusting for previously known covariates.

**Results:** Of 102 close family members (mean age 52 ± 15 years, 70% female, 40% non-Hispanic white, 21% Black, 33% Hispanic/Latinx, 22% with pre-existing psychiatric illness) with complete data, the mean PHQ-8 total score at a median duration of 28.5 days (interquartile range 10-63 days) from cardiac arrest was 7± 6, and the mean MSPSS score was 69 ± 15. Lower perceived social support was significantly associated with elevated levels of depressive symptoms in univariate (β=–0.11; p<0.01) and after adjusting for age, sex, race/ethnicity, and previous psychiatric history (β=–0.11; p<0.01). Similar inverse associations were seen with 1 month PTSD and generalized anxiety symptoms as secondary outcomes.

**Conclusions:** Close family members of cardiac arrest survivors’ perception of poor social support during hospitalization is associated with increased levels of depressive symptoms at 1 month. Longitudinal studies understanding the temporal associations between social support and psychological distress are warranted.

## INTRODUCTION

The subjective experience of cardiac arrest is deeply distressing and full of uncertainty, not only for patients themselves but also for their close family members. Our scoping review showed that one year after cardiac arrest, 14% of family members of cardiac arrest survivors report depression, much higher than the general population’s prevalence of 8.3%, with similarly elevated findings for post-traumatic stress disorder (PTSD) and anxiety [1-3]. Indeed, psychological distress is even more prevalent among family members than among cardiac arrest survivors themselves [4]. Despite this, there is very little known about the prevalence and predictors of distress in family members of cardiac arrest survivors [3].

In studies of family members’ mental health following their loved one’s illness, perceived social support has long been understood to be a key protective factor. Previous research has found that low perceived social support is associated with worse mental health outcomes in family members of prolonged mechanical ventilation, traumatic brain injury, and spinal cord injury patients [5-7], but this association has never been quantified in family members of cardiac arrest survivors [8,9].

In a consecutive sample of cardiac arrest patients’ family members, we assessed perceived social support and depressive symptom burden. Our objectives were: (1) to estimate the prevalence of indicators of psychological distress; 2) to estimate the correlation between perceived social support and psychological distress among family members; and (3) to describe the social support networks of family members in the early weeks following cardiac arrest.

## METHODS

### Study design

The Cardiac Arrest Neuropsychological Outcomes Evaluation (CANOE) cross-sectional study (N=152) was conducted at Columbia University Irving Medical Center (CUIMC) from August 16, 2021 to June 28, 2023. Close family members of consecutive cardiac arrest patients hospitalized at CUIMC completed a questionnaire at hospital discharge after the cardiac arrest. The questionnaire was completed either on paper or through Research Electronic Data Capture (REDCap), a secure online survey platform. This study protocol was approved by the Columbia University Institutional Review Board.

### Study participants

Inclusion criteria were: (1) age 18 years or older, (2) close family members of a cardiac arrest patient at CUIMC, and (3) English- or Spanish-speaking. Exclusion criteria were: (1) limited English or Spanish fluency, (2) cardiac arrest patients without a close family member at the bedside, and (3) bereavement before the time of approach by a study member.

### Questionnaire development and measures

We collected self-reported demographic information from participants, and clinical characteristics of the patients were retrieved from medical charts. (Table 1).

**Table 1.**
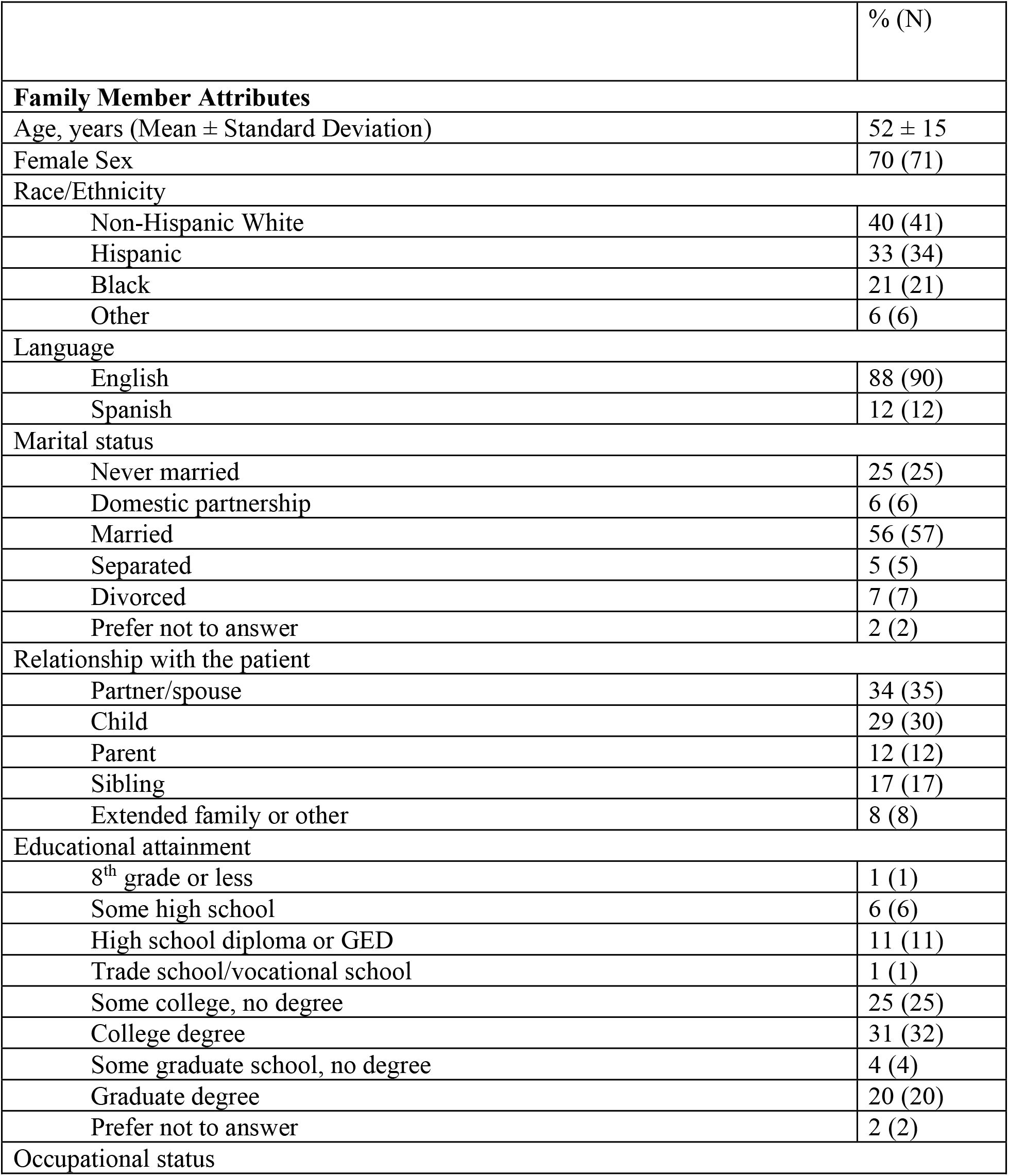
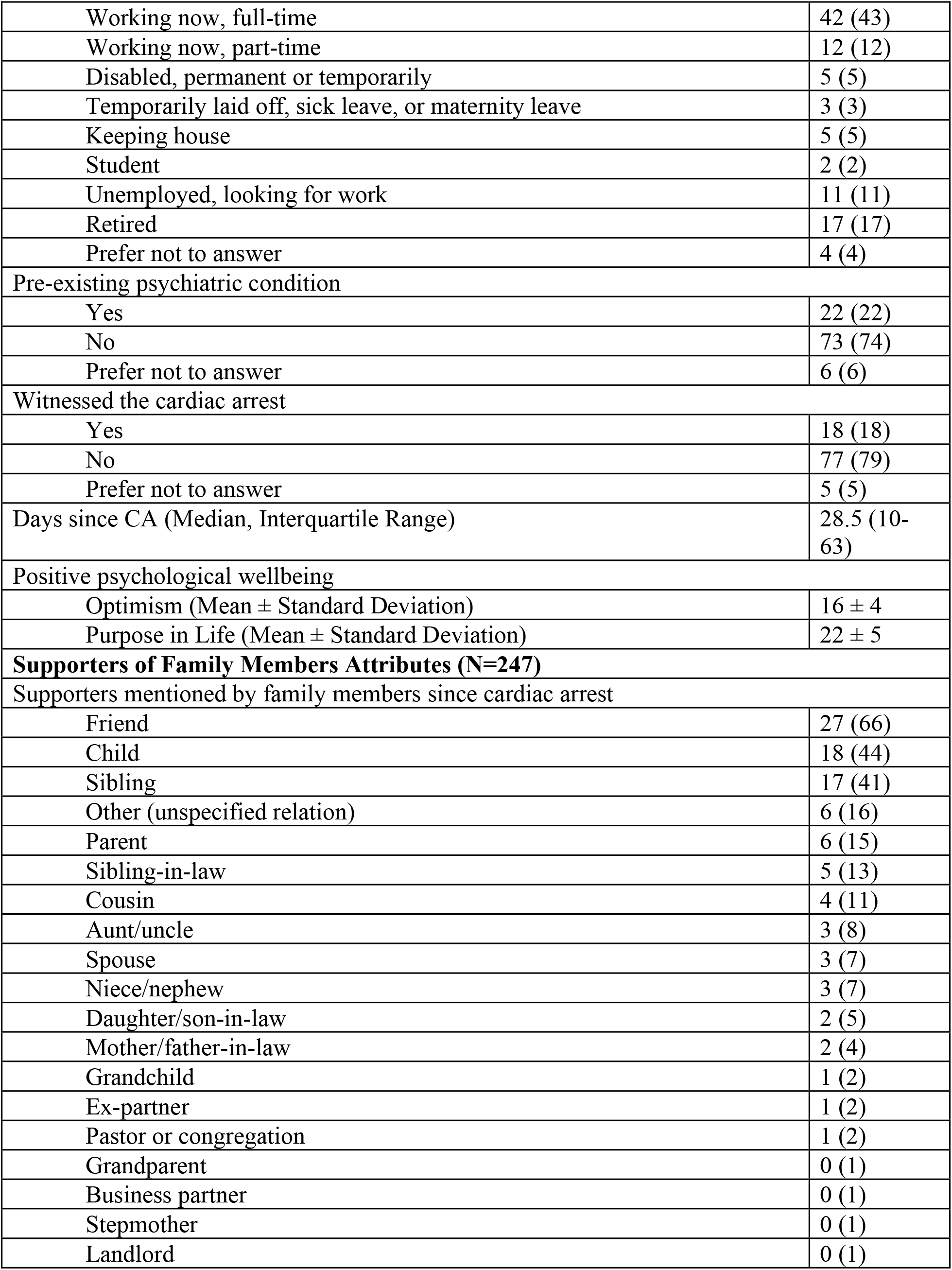
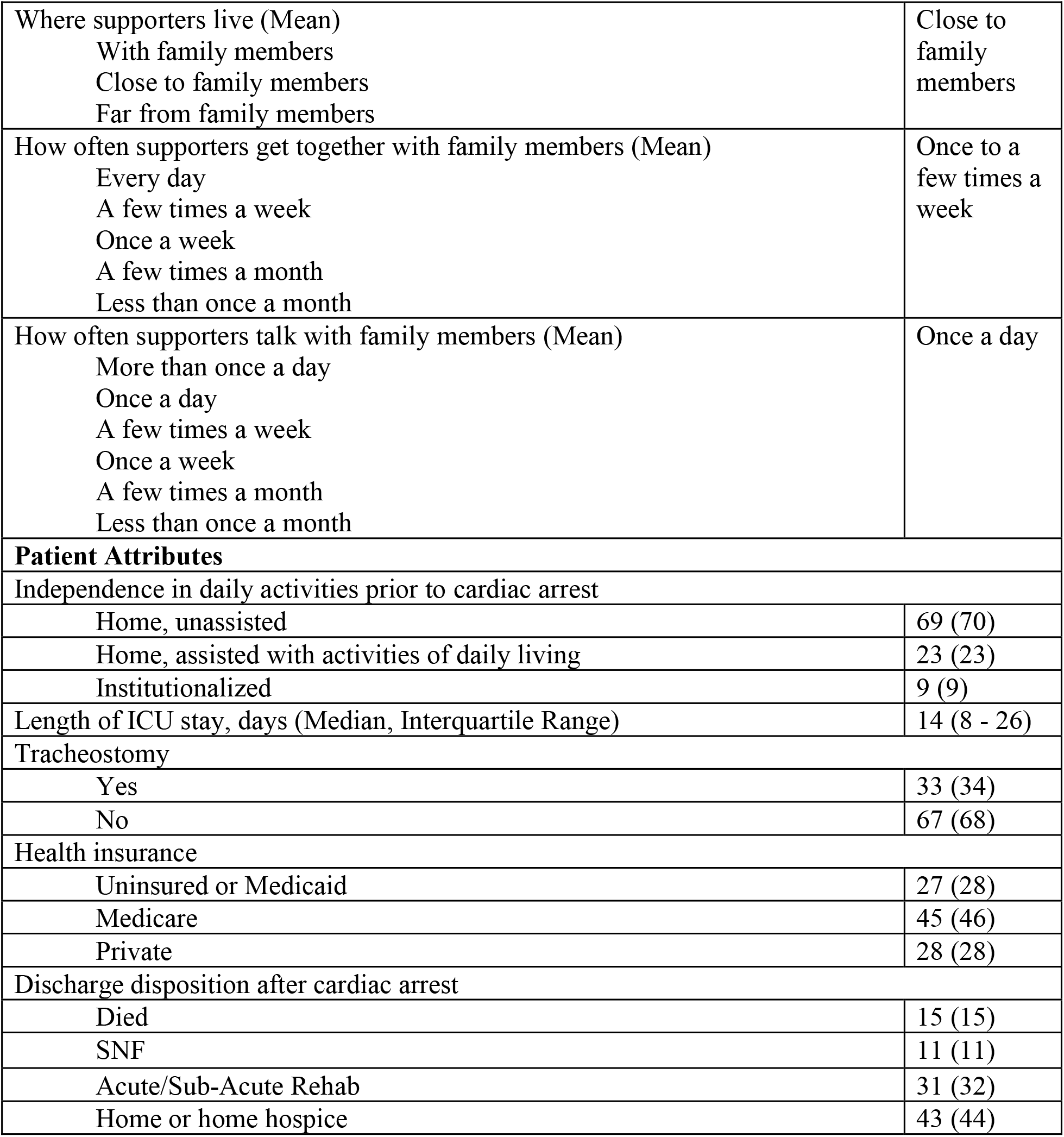
Characteristics of Family Members of Cardiac Arrest Survivors (N=102)

|  | % (N) |
| --- | --- |
| **Family Member Attributes** |  |
| Age, years (Mean ± Standard Deviation) | 52 ± 15 |
| Female Sex | 70 (71) |
| Race/Ethnicity |  |
| Non-Hispanic White | 40 (41) |
| Hispanic | 33 (34) |
| Black | 21 (21) |
| Other | 6 (6) |
| Language |  |
| English | 88 (90) |
| Spanish | 12 (12) |
| Marital status |  |
| Never married | 25 (25) |
| Domestic partnership | 6 (6) |
| Married | 56 (57) |
| Separated | 5 (5) |
| Divorced | 7 (7) |
| Prefer not to answer | 2 (2) |
| Relationship with the patient |  |
| Partner/spouse | 34 (35) |
| Child | 29 (30) |
| Parent | 12 (12) |
| Sibling | 17 (17) |
| Extended family or other | 8 (8) |
| Educational attainment |  |
| 8<sup>th</sup> grade or less | 1 (1) |
| Some high school | 6 (6) |
| High school diploma or GED | 11 (11) |
| Trade school/vocational school | 1 (1) |
| Some college, no degree | 25 (25) |
| College degree | 31 (32) |
| Some graduate school, no degree | 4 (4) |
| Graduate degree | 20 (20) |
| Prefer not to answer | 2 (2) |
| Occupational status |  |
| Working now, full-time | 42 (43) |
| Working now, part-time | 12 (12) |
| Disabled, permanent or temporarily | 5 (5) |
| Temporarily laid off, sick leave, or maternity leave | 3 (3) |
| Keeping house | 5 (5) |
| Student | 2 (2) |
| Unemployed, looking for work | 11 (11) |
| Retired | 17 (17) |
| Prefer not to answer | 4 (4) |
| Pre-existing psychiatric condition |  |
| Yes | 22 (22) |
| No | 73 (74) |
| Prefer not to answer | 6 (6) |
| Witnessed the cardiac arrest |  |
| Yes | 18 (18) |
| No | 77 (79) |
| Prefer not to answer | 5 (5) |
| Days since CA (Median, Interquartile Range) | 28.5 (10-63) |
| Positive psychological wellbeing |  |
| Optimism (Mean ± Standard Deviation) | 16 ± 4 |
| Purpose in Life (Mean ± Standard Deviation) | 22 ± 5 |
| **Supporters of Family Members Attributes (N=247)** |  |
| Supporters mentioned by family members since cardiac arrest |  |
| Friend | 27 (66) |
| Child | 18 (44) |
| Sibling | 17 (41) |
| Other (unspecified relation) | 6 (16) |
| Parent | 6 (15) |
| Sibling-in-law | 5 (13) |
| Cousin | 4 (11) |
| Aunt/uncle | 3 (8) |
| Spouse | 3 (7) |
| Niece/nephew | 3 (7) |
| Daughter/son-in-law | 2 (5) |
| Mother/father-in-law | 2 (4) |
| Grandchild | 1 (2) |
| Ex-partner | 1 (2) |
| Pastor or congregation | 1 (2) |
| Grandparent | 0 (1) |
| Business partner | 0 (1) |
| Stepmother | 0 (1) |
| Landlord | 0 (1) |
| Where supporters live (Mean)<br>With family members<br>Close to family members<br>Far from family members | Close to family members |
| How often supporters get together with family members (Mean)<br>Every day<br>A few times a week<br>Once a week<br>A few times a month<br>Less than once a month | Once to a few times a week |
| How often supporters talk with family members (Mean)<br>More than once a day<br>Once a day<br>A few times a week<br>Once a week<br>A few times a month<br>Less than once a month | Once a day |
| **Patient Attributes** |  |
| Independence in daily activities prior to cardiac arrest |  |
| Home, unassisted | 69 (70) |
| Home, assisted with activities of daily living | 23 (23) |
| Institutionalized | 9 (9) |
| Length of ICU stay, days (Median, Interquartile Range) | 14 (8 - 26) |
| Tracheostomy |  |
| Yes | 33 (34) |
| No | 67 (68) |
| Health insurance |  |
| Uninsured or Medicaid | 27 (28) |
| Medicare | 45 (46) |
| Private | 28 (28) |
| Discharge disposition after cardiac arrest |  |
| Died | 15 (15) |
| SNF | 11 (11) |
| Acute/Sub-Acute Rehab | 31 (32) |
| Home or home hospice | 43 (44) |

#### Psychological Distress

Our primary outcome, depressive symptom burden, was measured using the Patient Health Questionnaire-8 (PHQ-8), an 8-item self-report instrument with each item rated on a scale of 0 to 3 (“Not at all,” “Several days,” “More than half the days,” “Nearly every day”). A cutoff of 10 or higher indicates major depression [10].

Our secondary outcomes, post-traumatic stress disorder, and generalized anxiety disorder symptoms, were measured using the PTSD Checklist for DSM-5 (PCL-5) [11] and Generalized Anxiety Disorder 2-item (GAD-2) [12].

#### Perceived Social Support

We used a 12-item questionnaire assessing an individual’s perceived level of social support. It includes three subscales for support from family, friends, or significant others. Each item is rated on a scale of 1 (“very strongly disagree”) to 7 (“very strongly agree”), and a cutoff of 61 or higher indicates high perceived social support [13].

Further, to understand the social support network, participants also listed up to five people who have supported them since the cardiac arrest, their relation, physical proximity, how often they get together, and how often they talk.

### Statistical analysis

Using the PHQ-8 score to categorize all participants as meeting major depression criteria (≥10) versus not, we identified statistically significant characteristics between the two groups using chi-squared and two-sample T tests.

We next performed univariate and multivariate linear regression analysis including these characteristics to identify factors significantly associated with PHQ-8 score. Although language and occupational status were significantly associated with major depression, we chose to exclude them from the multivariate models given the small proportions of participants in each group (Table 2). We additionally included sex as a demographic factor *a priori* given previous research showing that women experience significantly worse psychiatric outcomes at hospital discharge than men [14]. We also included previous psychiatric history given its previously established association with cardiac arrest-induced psychological distress [15]. <u>Multivariate model 1</u>, in addition to total MSPSS score, adjusts for family member demographics (age, sex, race/ethnicity). <u>Multivariate model 2</u> includes model 1 and adds previous psychiatric history. All statistical analysis was performed using STATA 18 and a p-value of <0.05 to indicate significance.

**Table 2.**
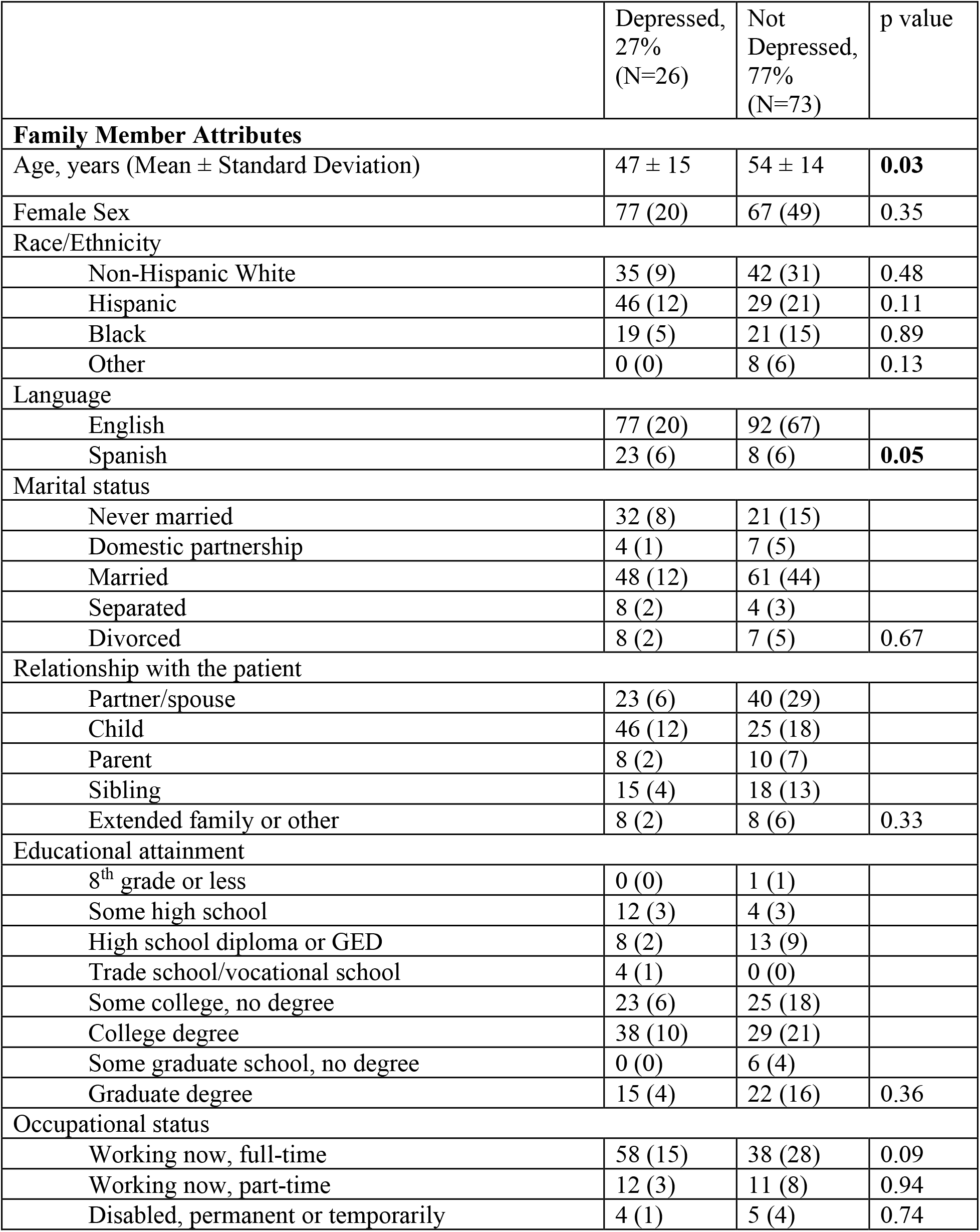
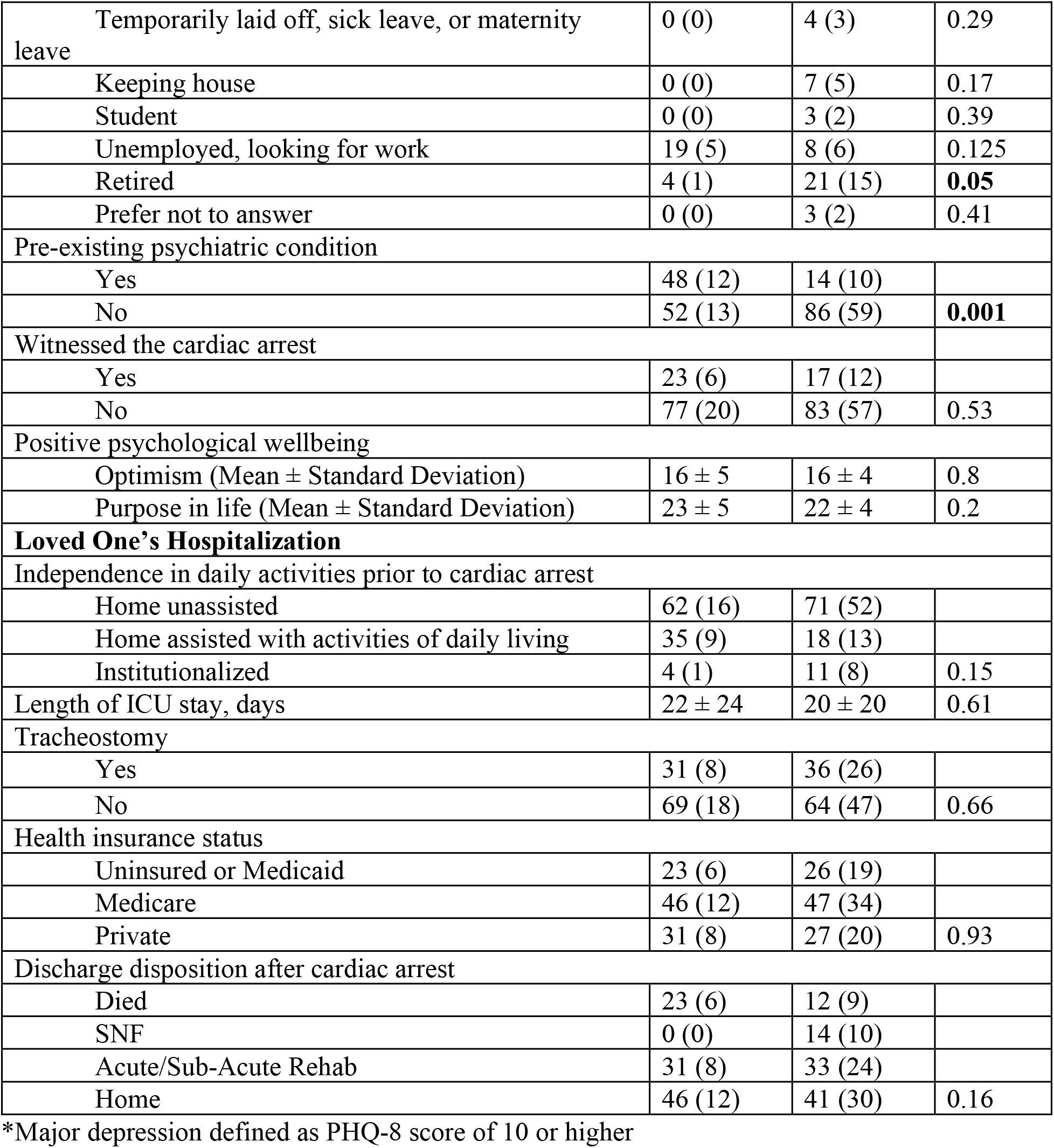
Comparing Characteristics of Family Members by Depression Status*.

|  | Depressed,<br>27%<br>(N=26) | Not<br>Depressed,<br>77%<br>(N=73) | p value |
| --- | --- | --- | --- |
| **Family Member Attributes** |  |  |  |
| Age, years (Mean ± Standard Deviation) | 47 ± 15 | 54 ± 14 | **0.03** |
| Female Sex | 77 (20) | 67 (49) | 0.35 |
| Race/Ethnicity |  |  |  |
| Non-Hispanic White | 35 (9) | 42 (31) | 0.48 |
| Hispanic | 46 (12) | 29 (21) | 0.11 |
| Black | 19 (5) | 21 (15) | 0.89 |
| Other | 0 (0) | 8 (6) | 0.13 |
| Language |  |  |  |
| English | 77 (20) | 92 (67) |  |
| Spanish | 23 (6) | 8 (6) | **0.05** |
| Marital status |  |  |  |
| Never married | 32 (8) | 21 (15) |  |
| Domestic partnership | 4 (1) | 7 (5) |  |
| Married | 48 (12) | 61 (44) |  |
| Separated | 8 (2) | 4 (3) |  |
| Divorced | 8 (2) | 7 (5) | 0.67 |
| Relationship with the patient |  |  |  |
| Partner/spouse | 23 (6) | 40 (29) |  |
| Child | 46 (12) | 25 (18) |  |
| Parent | 8 (2) | 10 (7) |  |
| Sibling | 15 (4) | 18 (13) |  |
| Extended family or other | 8 (2) | 8 (6) | 0.33 |
| Educational attainment |  |  |  |
| 8<sup>th</sup> grade or less | 0 (0) | 1 (1) |  |
| Some high school | 12 (3) | 4 (3) |  |
| High school diploma or GED | 8 (2) | 13 (9) |  |
| Trade school/vocational school | 4 (1) | 0 (0) |  |
| Some college, no degree | 23 (6) | 25 (18) |  |
| College degree | 38 (10) | 29 (21) |  |
| Some graduate school, no degree | 0 (0) | 6 (4) |  |
| Graduate degree | 15 (4) | 22 (16) | 0.36 |
| Occupational status |  |  |  |
| Working now, full-time | 58 (15) | 38 (28) | 0.09 |
| Working now, part-time | 12 (3) | 11 (8) | 0.94 |
| Disabled, permanent or temporarily | 4 (1) | 5 (4) | 0.74 |
| Temporarily laid off, sick leave, or maternity leave | 0 (0) | 4 (3) | 0.29 |
| Keeping house | 0 (0) | 7 (5) | 0.17 |
| Student | 0 (0) | 3 (2) | 0.39 |
| Unemployed, looking for work | 19 (5) | 8 (6) | 0.125 |
| Retired | 4 (1) | 21 (15) | **0.05** |
| Prefer not to answer | 0 (0) | 3 (2) | 0.41 |
| Pre-existing psychiatric condition |  |  |  |
| Yes | 48 (12) | 14 (10) |  |
| No | 52 (13) | 86 (59) | **0.001** |
| Witnessed the cardiac arrest |  |  |  |
| Yes | 23 (6) | 17 (12) |  |
| No | 77 (20) | 83 (57) | 0.53 |
| Positive psychological wellbeing |  |  |  |
| Optimism (Mean ± Standard Deviation) | 16 ± 5 | 16 ± 4 | 0.8 |
| Purpose in life (Mean ± Standard Deviation) | 23 ± 5 | 22 ± 4 | 0.2 |
| **Loved One's Hospitalization** |  |  |  |
| Independence in daily activities prior to cardiac arrest |  |  |  |
| Home unassisted | 62 (16) | 71 (52) |  |
| Home assisted with activities of daily living | 35 (9) | 18 (13) |  |
| Institutionalized | 4 (1) | 11 (8) | 0.15 |
| Length of ICU stay, days | 22 ± 24 | 20 ± 20 | 0.61 |
| Tracheostomy |  |  |  |
| Yes | 31 (8) | 36 (26) |  |
| No | 69 (18) | 64 (47) | 0.66 |
| Health insurance status |  |  |  |
| Uninsured or Medicaid | 23 (6) | 26 (19) |  |
| Medicare | 46 (12) | 47 (34) |  |
| Private | 31 (8) | 27 (20) | 0.93 |
| Discharge disposition after cardiac arrest |  |  |  |
| Died | 23 (6) | 12 (9) |  |
| SNF | 0 (0) | 14 (10) |  |
| Acute/Sub-Acute Rehab | 31 (8) | 33 (24) |  |
| Home | 46 (12) | 41 (30) | 0.16 |

\*Major depression defined as PHQ-8 score of 10 or higher

## RESULTS

In total, 102 family members with complete data were included. Of 438 non-fatal cardiac arrests admitted to CUIMC, 247 were excluded due to bereavement during hospitalization, limited English/Spanish fluency, or lack of a close family member. Fifty-four family members declined to participate, and 35 withdrew or were lost to follow-up (Supplementary Figure 1). Characteristics of this cohort are shown in Table 1. The average age of family members was 52 years. Seventy-one (70%) were female, 61 (60%) identified as part of a minority racial/ethnic group, 12 (12%) were Spanish-speaking, 35 (34%) were the patient’s spouse or partner, 55 (54%) were working part- or full-time, 22 (22%) reported pre-existing psychiatric conditions, and 18 (18%) witnessed the cardiac arrest. Family members completed the questionnaire a median of 28.5 days (interquartile range 10-63 days) after the cardiac arrest.

Participants’ loved ones stayed in the intensive care unit (ICU) for a median of 14 days. Before cardiac arrest, 93 (92%) lived at home. Following cardiac arrest, 34 (33%) underwent tracheostomy, and 44 (43%) were ultimately discharged home. Additionally, 28 (27%) were uninsured or on Medicaid.

### Perceived social support and depression

Twenty-six participants (27%) met criteria for major depression. Participants who met criteria for major depression were more likely to be younger, Spanish-speaking, non-retired, and have previous psychiatric history (Table 2). No differences in depression status were found based on race/ethnicity, positive psychological well-being, or patient characteristics.

In our univariate analysis, higher PHQ-8 scores were significantly associated with lower total MSPSS scores (β =-0.11, p=0.003) and previous psychiatric history (β =4.49, p<0.001). In our multivariate model, lower total MSPSS scores, younger age, female sex, and previous psychiatric history were significantly associated with higher PHQ-8 scores (Table 3). Notably, the family (β=-0.33, p=0.001), friends (β =-0.18, p=0.037), and significant other (β =-0.27, p=0.003) subscales of the MSPSS all passed the threshold of significance when controlling for demographics and previous psychiatric history, suggesting that all three subscales drive the association shown here between total MSPSS and PHQ-8 scores.

**Table 3.**
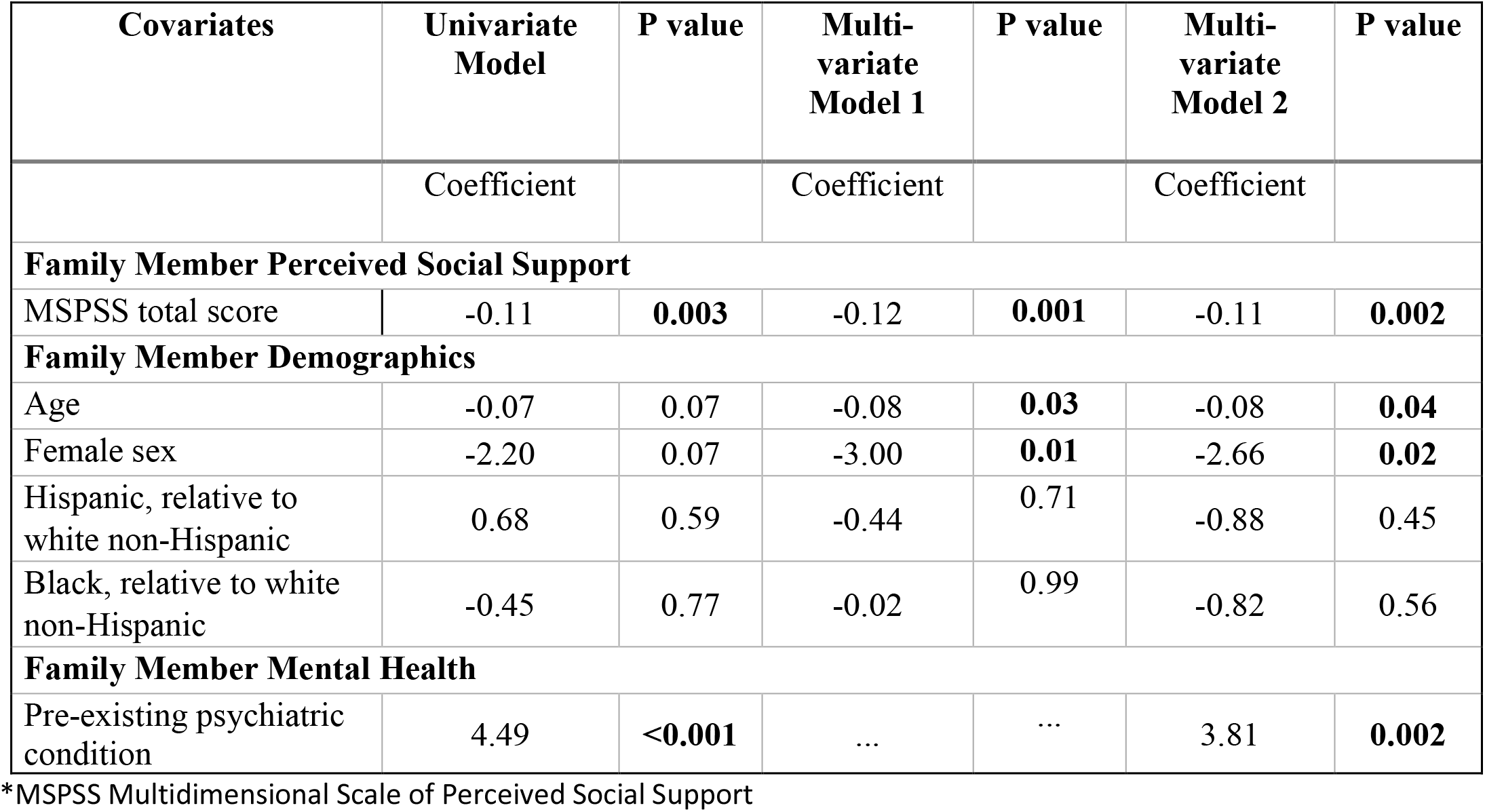
Associations Between Family Member Characteristics and Total PHQ-8 Score.

| **Covariates** | **Univariate Model** | **P value** | **Multi-variate Model 1** | **P value** | **Multi-variate Model 2** | **P value** |
| --- | --- | --- | --- | --- | --- | --- |
|  | Coefficient |  | Coefficient |  | Coefficient |  |
| **Family Member Perceived Social Support** |  |  |  |  |  |  |
| MSPSS total score | -0.11 | **0.003** | -0.12 | **0.001** | -0.11 | **0.002** |
| **Family Member Demographics** |  |  |  |  |  |  |
| Age | -0.07 | 0.07 | -0.08 | **0.03** | -0.08 | **0.04** |
| Female sex | -2.20 | 0.07 | -3.00 | **0.01** | -2.66 | **0.02** |
| Hispanic, relative to white non-Hispanic | 0.68 | 0.59 | -0.44 | 0.71 | -0.88 | 0.45 |
| Black, relative to white non-Hispanic | -0.45 | 0.77 | -0.02 | 0.99 | -0.82 | 0.56 |
| **Family Member Mental Health** |  |  |  |  |  |  |
| Pre-existing psychiatric condition | 4.49 | **<0.001** | ... | ... | 3.81 | **0.002** |

\*MSPSS Multidimensional Scale of Perceived Social Support

Having low or moderate perceived social support (MSPSS < 61) was also significantly associated with higher PHQ-8 (β=4.32, p=0.001).

### Perceived social support and PTSD

Similarly, lower total MSPSS scores were significantly associated with higher PCL-5 (β=-0.30, p<0.001) scores. PCL-5 scores were significantly associated with the family (β=-0.73, p=0.003), friends (β=-0.61, p=0.003), and significant other (β=-0.71, p=0.001) subscales of the MSPSS,. Much like the PHQ-8, having low or moderate perceived social support (MSPSS < 61) was significantly associated with higher PCL-5 scores (β=9.07, p=0.001).

### Perceived social support and generalized anxiety

Finally, lower total MSPSS scores were significantly associated with higher GAD-2 (β=-0.04, p=0.01) scores. GAD-2 scores were significantly associated with the family (β=-0.11, p=0.008) and significant other (β=-0.08, p=0.03) subscales of the MSPSS, but not the friends subscale (β=-0.06, p=0.07). Having low or moderate perceived social support (MSPSS < 61) was significantly associated with GAD-2 scores (β =1.28, p=0.01).

### Self-reported social support networks of close family members

Participants listed a wide range of sources of social support since their loved one’s cardiac arrest (Table 1). The most frequently mentioned type of supporter was friends (N=66, 27%), followed by children (N=44, 18%) and siblings (N=41, 17%). On average, participants reported that their supporters lived close by, met with them once to a few times a week, and talked with them once a day. PHQ-8 score was significantly negatively associated with frequency of meeting up (β=-1.13, p=0.02), but not physical proximity (β=-0.91, p=0.46) or frequency of talking (β=-0.78, p=0.21), suggesting that having supporters who can meet in-person frequently may be protective against depression.

## DISCUSSION

In this study, we found that close family members with higher perceived social support reported significantly lower burden of depressive symptoms. Our findings align with previous research on depressive outcomes in family members of ICU patients, which also found that younger age and lower perceived social support are significantly associated with worse mental health outcomes, while patient characteristics are not [5]. However, while previous research has found that spouses and family members who witnessed resuscitative efforts after cardiac arrest have higher depressive symptom burden, we did not find a significant association between witness status and depression [16]. We also did not find that spouses had significantly more depressive symptoms compared to other close family members.

Our subscale analysis found that support from family, friends, and significant others all drive the association between perceived social support and depression, suggesting that all three types of supporters matter for family member mental health in the first few weeks after cardiac arrest. It is notable that participants’ definitions of social support appear to go beyond these three categories; when prompted to list supporters, participants also mentioned other types of supporters, such as their religious communities and former spouses. Listed supporters tended to be individuals that participants lived near, talked to, and met up with frequently, suggesting future interventions targeting family member-supporter dyads may need to consider proximity and accessibility when selecting supporters.

Our findings are of particularly urgent importance given that 27% of our participants met criteria for major depression, which is higher than among cardiac arrest survivors and more than triple the prevalence among the general population [2, 17]. Notably, this is a much higher prevalence of depression than found in previous studies of out-of-hospital cardiac arrest patients’ relatives 90 days after admission [16]. Given that other research has found that 16% of caregivers of ICU patients do not experience a decrease in depressive symptom burden with time, identifying and intervening for vulnerable family members early is of great importance [5]. Additionally, given that participants who were female, younger, or previously diagnosed with psychiatric conditions reported significantly higher depressive symptom burden, these family members may be in especially critical need of interventions targeting perceived social support. We found no significant differences across race/ethnicity, suggesting future social support-based interventions may benefit family members of all race/ethnicity groups equally.

### Study strengths

This study has several notable strengths. To our knowledge, this study recruited the largest sample size of cardiac arrest patients’ family members to date. Our sample includes a diverse group of family members, with ages ranging from 18-80 years, a wide race/ethnicity distribution, and both in-hospital and out-of-hospital cardiac arrests. This speaks to the high generalizability of our findings.

This is also the first study, to our knowledge, to examine the role of perceived social support in depressive outcomes with a focus on family members of cardiac arrest patients. These findings help fill the gap in the literature, which predominantly features cardiac arrest patient outcomes.

### Study limitations and next steps

There are also some limitations to this study. First, given the cross-sectional nature of this study, the results reported here cannot be extrapolated to long-term outcomes of family members. In the future, we plan to follow up with participants at several time points to assess changes in perceived social support and depressive symptom burden over time to better understand their needs after cardiac arrest.

Additionally, our characterization of family members’ social support networks is limited by only allowing participants to report up to five people who have supported them since the cardiac arrest. To fully map social support networks in the future, we plan to prompt participants to provide an exhaustive list of all supporters, the nature of their relationships, and the strength of their relationships. Previous research has demonstrated that detailed social network data can be used to develop social interventions, like network interventions and social skills training, to improve physical and mental health [18, 19]. A more complete understanding of the key individuals composing family members’ support networks is essential for building future interventions.

Prior studies have highlighted the need to optimize ways to improve family members’ perceived social support after life-threatening illnesses [20]. The next step for this work will be to follow family members in a longitudinal study assessing perceived social support and psychological distress at multiple time points after cardiac arrest.

## Data Availability

All data produced in the present study are available upon reasonable request to the authors.

## SOURCES OF FUNDING

Sachin Agarwal was a principal investigator on a related NIH grant (R01-HL153311) that provided salary support for his effort and funded the current study. Christine DeForge was supported by an institutional training grant funded by the National Institutes of Health/National Center for Advancing Translational Sciences (TL1TR001875). Mina Yuan was supported by an institutional training grant funded by the NIH (5T35HL007616).

## DISCLOSURES

Authors have nothing to disclose.

---

**Supplementary Figure 1.**
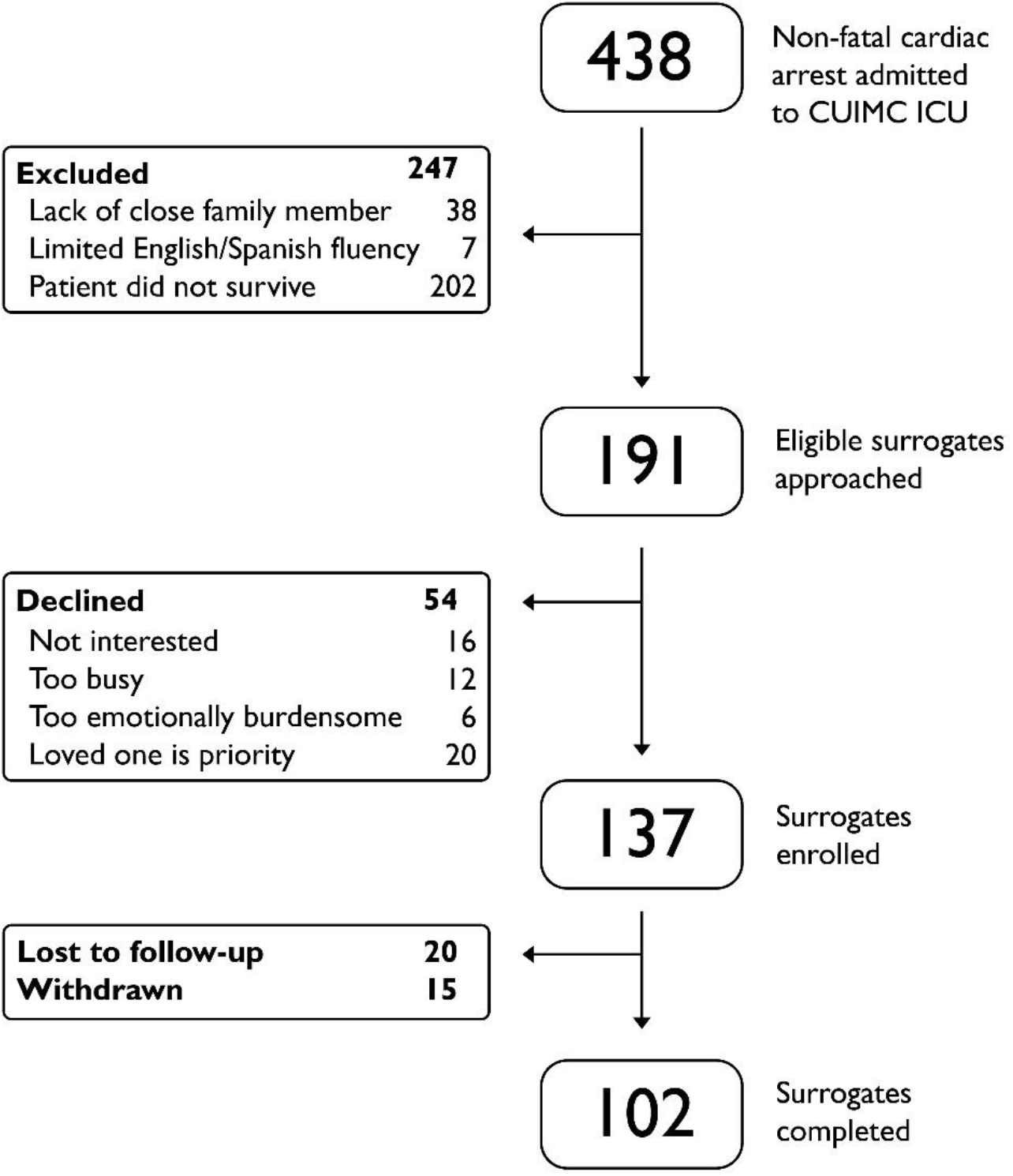
CONSORT Diagram.

**Supplementary Table 1.** Associations Between Family Member Characteristics and PTSD (total PCL score)

| Covariates | Univariate Model | P value | Multi-variate Model 1 | P value | Multi-variate Model 2 | P value |
| --- | --- | --- | --- | --- | --- | --- |
|  | Coefficient |  | Coefficient |  | Coefficient |  |
| **Family Member Perceived Social Support** |  |  |  |  |  |  |
| MSPSS total score | -0.32 | **<0.001** | -0.33 | **<0.001** | -0.30 | **<0.001** |
| **Family Member Demographics** |  |  |  |  |  |  |
| Age | -0.07 | 0.43 | -0.09 | 0.28 | -0.07 | 0.45 |
| Female sex | -1.14 | 0.67 | -2.02 | 0.43 | -1.24 | 0.63 |
| Hispanic, relative to white non-Hispanic | 3.14 | 0.26 | 1.67 | 0.54 | 0.68 | 0.80 |
| Black, relative to white non-Hispanic | 2.15 | 0.53 | 2.04 | 0.53 | 0.09 | 0.98 |
| **Family Member Mental Health** |  |  |  |  |  |  |
| Pre-existing psychiatric condition | 9.25 | **0.002** | ... | ... | 7.86 | **0.01** |

**Supplementary Table 2.** Associations Between Family Member Characteristics and Generalized Anxiety (total GAD-2 Score)

| Covariates | Univariate Model | P value | Multi-variate Model 1 | P value | Multi-variate Model 2 | P value |
| --- | --- | --- | --- | --- | --- | --- |
|  | Coefficient |  | Coefficient |  | Coefficient |  |
| **Family Member Perceived Social Support** |  |  |  |  |  |  |
| MSPSS total score | -0.03 | **0.02** | -0.04 | **0.01** | -0.04 | **0.01** |
| **Family Member Demographics** |  |  |  |  |  |  |
| Age | -0.03 | 0.02 | -0.04 | **0.01** | -0.04 | **0.01** |
| Female sex | -0.44 | 0.35 | -0.74 | 0.11 | -0.79 | 0.09 |
| Hispanic, relative to white non-Hispanic | 0.28 | 0.56 | -0.13 | 0.79 | -0.41 | 0.39 |
| Black, relative to white non-Hispanic | -0.09 | 0.87 | 0.05 | 0.92 | -0.20 | 0.73 |

| **Family Member Mental Health** |  |  |  |  |  |  |
| --- | --- | --- | --- | --- | --- | --- |
| Pre-existing psychiatric condition | 1.37 | **0.01** | ... | ... | 1.07 | **0.03** |

## Notes

### Competing Interest Statement

The authors have declared no competing interest.

### Author Declarations

IRB of Columbia University Medical Center gave ethical approval for this work.

